# The COVID-19 mortality effects of underlying health conditions in India: a modelling study

**DOI:** 10.1101/2020.07.05.20140343

**Authors:** Paul Novosad, Radhika Jain, Alison Campion, Sam Asher

## Abstract

**Objective:** To model how known COVID-19 comorbidities will affect mortality rates and the age distribution of mortality in a large lower middle income country (India), as compared with a high income country (England), and to identify which health conditions drive any differences.

**Design:** Modelling study.

**Setting:** England and India.

**Participants:** 1,375,548 respondents aged 18 to 99 to the District Level Household Survey-4 and Annual Health Survey in India. Additional information on health condition prevalence on individuals aged 18 to 99 was obtained from the Health Survey for England and the Global Burden of Diseases, Risk Factors, and Injuries Studies (GBD).

**Main outcome measures:** The primary outcome was the proportional increase in age-specific mortality in each country due to the prevalence of each COVID-19 mortality risk factor (diabetes, hypertension, obesity, chronic heart disease, respiratory illness, kidney disease, liver disease, and cancer, among others). The combined change in overall mortality and the share of deaths under 60 from the combination of risk factors was estimated in each country.

**Results:** Relative to England, Indians have higher rates of diabetes (10.6% vs. 8.5%), chronic respiratory disease (4.8% vs. 2.5%), and kidney disease (9.7% vs. 5.6%), and lower rates of obesity (4.4% vs. 27.9%), chronic heart disease (4.4% vs. 5.9%), and cancer (0.3% vs. 2.8%). Population COVID-19 mortality in India relative to England is most increased by diabetes (+5.4%) and chronic respiratory disease (+2.3%), and most reduced by obesity (−9.7%), cancer (−3.2%), and chronic heart disease (−1.9%). Overall, comorbidities lower mortality in India relative to England by 9.7%. Accounting for demographics and population health explains a third of the difference in share of deaths under age 60 between the two countries.

**Conclusions:** Known COVID-19 health risk factors are not expected to have a large effect on aggregate mortality or its age distribution in India relative to England. The high share of COVID-19 deaths from people under 60 in low- and middle-income countries (LMICs) remains unexplained. Understanding mortality risk associated with health conditions prevalent in LMICs, such as malnutrition and HIV/AIDS, is essential for understanding differential mortality.

**SUMMARY BOX:** *What is already known on this topic:* COVID-19 infections in low- and middle-income countries (LMICs) are rising rapidly, with the burden of mortality concentrated at much younger ages than in rich countries. A range of pre-existing health conditions can increase the severity of COVID-19 infections. It is feared that poor population health may worsen the severity of the pandemic in LMICs.

*What this study adds:* The COVID-19 comorbidities that have been studied to date may have only a very small effect on aggregate mortality in India relative to England and do not shift the mortality burden toward lower ages at all. India’s younger demographics can explain only a third of the substantial difference in the share of deaths under age 60 between India and England. However, mortality risk associated with health conditions prevalent in LMICs, such as malnutrition and HIV/AIDS, is unknown and research on this topic is urgently needed.

## INTRODUCTION

As governments around the world ease social distancing measures imposed to limit the transmission of COVID-19, the number of global cases is rising. A growing share of cases is now coming from low- and middle-income countries (LMICs) in Asia, Africa, and the Americas that were largely spared in the initial stages of the pandemic.^1^ Because the severity of infection is substantially increasing with age, forecasts have projected much lower aggregate mortality rates in LMICs than in wealthier countries.^2–4^

However, reported fatality numbers from LMICs to date have suggested a much greater share of COVID-19 deaths among the young. 30.5% of deaths in Brazil and 50% of deaths in India have occurred in those under age 60; 27% of deaths in Mexico have occurred in those under 50.^5–7^ In contrast, individuals under 65 have accounted for only 5–13% of deaths in 10 European countries and Canada and 8–24% in US states.^8^ It is not presently known whether the different age pattern of deaths in LMICs is driven by erroneous reporting, differences in infection patterns, younger populations, or worse underlying population health.

Many modelling studies have presumed that worse population health in poor countries will lead to excess mortality, or else ignored differential population health as a factor entirely.^4,9,10^ To date there has been limited analysis of the prevalence in LMICs of the specific conditions associated with increased COVID-19 severity, such as diabetes, obesity, cardiovascular disease, hypertension, and chronic kidney disease, nor of how they change the expected level and age distribution of mortality.^4,11–14^ Some studies have adjusted mortality estimates for population comorbidities by treating all comorbidities as equivalent or by multiplying the mortality rate by a fixed amount to adjust for population health.^15–17^ One study combined condition-specific prevalence and hazard ratios from a sample of hospitalizations, but excluded obesity and uncontrolled diabetes, and did not examine mortality or the age distribution of mortality as outcomes.^18^

Using England as a benchmark, this study examines how comorbidities understood to increase COVID-19 mortality are likely to affect COVID-19 mortality rates in aggregate and across the age distribution in India, identifying the specific risk factors with the largest mortality effects. We further study the extent to which accounting for differences in demographics and underlying health conditions can explain the increased share of deaths among the young in India relative to England.

Our analysis focuses on India and on the COVID-19 risk factors that are currently documented. India presently has the third highest number of cumulative COVID-19 infections in the world and the highest growth rate in infections of any major country, making it an essential population to study.^1^ The methodology is readily adjusted to account for new risk factors or data from other countries and may be useful for modelling the epidemic in a range of LMICs.

## METHODS

Our approach requires three types of data: (i) the relative risk of COVID-19 mortality associated with gender, age, and each health condition; (ii) the age-specific prevalence of each health condition in England and India; and (iii) the age and gender distributions for the two countries.

### Estimates of Relative Risk of COVID-19 Mortality from Comorbidities

We obtained estimates of COVID-19 mortality risk for a wide range of comorbidities from the OpenSAFELY study, a closed cohort study of 17,425,445 adults from England.^19^ This was the largest analysis of comorbidities associated with COVID-19 mortality to date and one of the few studies that estimates risk factors in a multivariate model adjusting for age, sex, and other health conditions. This adjustment is important because many COVID-19 comorbidities are increasing in age and their hazard ratios are thus biased upward in analyses not adjusting for age.

The OpenSAFELY study enrolled all individuals registered with a general practice within The Phoenix Partnership system on 1st February 2020, were 18 years or older upon enrolment, had at least one year of prior medical history within the system, and had recorded age and sex. The underlying dataset represents 40% of the population of England and the prevalence of health conditions in the study cohort is similar to estimates of population prevalence in England (appendix p 4). Patients were followed through 25th April. The outcome was in-hospital death among people with confirmed COVID-19 infections. Hazard Ratios (HRs) for mortality from a Cox proportional hazards model were estimated for a comprehensive list of risk factors described in other studies, adjusted for sex, age, and all other risk factors. As patient-level data from OpenSAFELY are not publicly available, we extracted HRs from the paper reporting results of the analysis.^19^

Ideally, HRs would measure mortality risk conditional upon infection, rather than on clinic attendance (this paper) or hospitalization (as in prior work).^18,20^ The HRs in this study reflect combined mortality and infection risk; the analysis assumes that pre-existing health conditions are not significant predictors of infection. However, condition HRs measuring mortality risk conditional upon hospitalization are similar to those used here.^11^

### Demographics and Risk Factor Prevalence in India and England

Age distributions and age-specific sex ratios for India and England were obtained from official censuses.

We obtained data on age-specific prevalence of health risk factors for India and England from multiple sources, prioritizing biomarker data where available, and matching definitions as closely as possible to the conditions for which HRs are available. We restrict samples to ages 18–99 for consistency with the HRs.

For India, we used biomarker data from two public population health surveys for obesity, diabetes, and hypertension. The fourth round of the Indian District Level Household Survey (DLHS-4) and the second round of the Annual Health Survey (AHS) were conducted between 2012 and 2014, jointly cover 94% of the Indian population, and provide the most recent nationwide direct measures of height, weight, fasting plasma glucose (FPG) and blood pressure (BP) for adults of all ages in India. Details of dataset construction are provided in the Appendix (p 1). For England, age-specific prevalences of obesity, hypertension, and diabetes were obtained from the nationally representative 2018 Health Survey for England, which collected symptoms and medical diagnoses for a range of illnesses, as well as direct measures of height, weight, blood pressure, and glycated haemoglobin (HbA1c).^21^

BMI was classified into no evidence of obesity (<30kg/m2), obese class one or two (30–39.9kg/m2), and obese class three (40+kg/m2). Hypertension was defined as systolic BP ≥140 mm Hg or diastolic BP ≥90 mm Hg (uncontrolled) or a medical diagnosis of hypertension with BP below the thresholds (controlled). Controlled and uncontrolled hypertension prevalence were reported separately but combined in the risk estimation for consistency with OpenSAFELY. OpenSAFELY classified controlled diabetes as Hba1c > 51 mmols/mol and Hba1c < 58mmols/mol, and uncontrolled diabetes as Hba1c ≥ 58 mmols/mol. Corresponding thresholds for the one-time FPG measures in the Indian dataset are not well defined. In England, prevalence was reported based on a threshold of HbA1c ≥ 48mmol/mol (6.5%). Therefore, we followed the standard screening and diagnosis thresholds recommended by the WHO and International Diabetes Federation and defined uncontrolled diabetes in India as a plasma glucose reading ≥126mg/dL [7.0mmol/L] if fasting or ≥200 mg/dL [11.1 mmol/L] if not fasting. We used the corresponding recommended threshold of HbA1c ≥48mmol/mol (6.5%) for uncontrolled diabetes in England.^22^ In both countries, we classified individuals with biomarkers below the thresholds but with a diagnosis of diabetes as having controlled diabetes.

Age-specific prevalence for asthma, chronic heart disease, kidney disease, stroke, dementia, haematological malignancies, and all other cancers were drawn from the Global Burden of Diseases, Risk Factors, and Injuries Studies (GBD) for India and England.^23^ OpenSAFELY reports separate HRs for cancers diagnosed <1 year ago, 1–4.9 years ago, and ≥5 years ago; because the year of diagnosis is unavailable in GBD, we used a single classification for each class of cancers and the HR for diagnosis <1 year ago. For chronic respiratory disease, we used COPD prevalence from the GBD for India and modelled COPD prevalence from the Clinical Practice Research Datalink cohort database for England.^24^ GBD prevalence of Parkinson’s disease, epilepsy, multiple sclerosis, and motor neuron disease were combined and classified as neurological disorders.

The following risk factors were not available for India and were excluded from the analysis for both England and India for comparability: fibrosing lung disease, bronchiectasis or cystic fibrosis, lupus, asthma with no recent OCS use, cancers diagnosed more than a year ago, organ transplant, and spleen disease. Given that the relationship between smoking and COVID-19 mortality remains under debate, we excluded it from the analysis.^25^ We also excluded ethnicity and socioeconomic status, which cannot be measured comparably across England and India and are unlikely to have similar relative risk in the two countries.

### Estimating the Contribution of Health Conditions to Population COVID-19 Mortality Risk

The OpenSAFELY study reports HRs for each age group, sex, and health condition with females 50–59 years with no conditions as the reference group.^19^ We transform the HR for each health condition *c* into a relative risk (*RR*_*c*_) assuming a population mortality rate *r* of 1%:

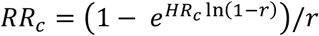

To obtain continuous relative risk for age, we used a polynomial interpolation for the log HR at each age, renormalizing with age 50 as the reference group (appendix p 5).

The increase in population mortality risk from a given health condition is increasing in the condition’s relative risk for COVID-19 mortality and its prevalence at each age. We defined the age- and condition-specific population relative risk *PRR*_*a, c*_ of condition *c* at age *a* as:

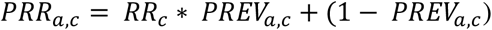

*PREV*_*a,c*_ is the prevalence of condition *c* at age *a. PRR*_*a,c*_ describes the proportional increase in mortality at age *a* driven by health condition *c*.

We combined PRRs to obtain an age-specific population relative risk of mortality arising from the combined prevalence of all of the health conditions :

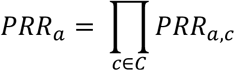

*PRR*_*a*_ isolates the expected mortality difference at each age between India and England that is driven by the combined prevalence of all of the health conditions studied. This approach implicitly assumes that the health conditions are uncorrelated with each other. Without microdata on the full set of health conditions, this assumption is unavoidable, but will bias the England vs. India comparison only if the correlation of health conditions is substantially different in the two countries. We explore the possible extent of this bias in the appendix. By using age-specific prevalence, our analysis fully accounts for the substantial correlations between age and health conditions.

We next calculated the increase in population mortality from each health condition across all ages, taking into account the age-specific prevalence of each health condition, its relative risk, and the population share at each age. The condition-specific population relative risk of each health condition across the full population (*PRR*_*c*_) is given by:

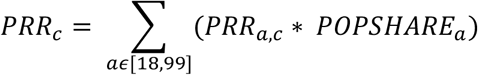

*PRR*_*c*_ is greater when the relative risk of condition *c* is higher, and when its prevalence is higher at ages with higher population and greater mortality risk. The combined effect on population mortality of all of the health conditions is given by the product of each condition-specific *PRR*_*c*_

Finally, we aggregated the population relative risks across health conditions in order to model the age distribution of deaths in each country. The number of deaths at each age *N*_*a*_ is the product of the mortality rate of the reference group (50-year-old women with no other risk factors), the population at age *a*, the age-specific population relative risk of the full set of health conditions, the PRR of gender, and the direct relative risk of COVID-19 mortality for an individual at age *a (RR*_*a*_):

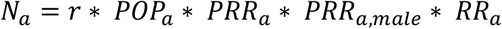

We plotted the age distribution of deaths as shares of all deaths rather than in levels, eliminating the need to assume a reference group mortality rate. We summarized the shape of the distribution by reporting the share of expected deaths in each country that are under the age of 60. We present results from three models: (i) England’s demographics and health distribution; (ii) India’s demographics and health distribution; and (iii) India’s demographics but England’s age-specific prevalence of health risk factors. The third model allowed us to examine the mortality shift that comes from differences in population health alone.

### Role of the Funding Source

The funder of the study had no role in study design, data collection, data analysis, data interpretation, or writing. The corresponding author had full access to the data in the study and had final responsibility for the decision to submit for publication.

### Patient and Public Involvement

Because this study uses existing epidemiological data, it was not appropriate to involve patients or the public in the research.

## RESULTS

### Prevalence of COVID-19 risk factors in India and England

Demographic characteristics and overall prevalence of risk factors are substantially different in India relative to England (table 1). 83.8% of Indian adults are below the age of 60, compared to 69.9% of English adults. Indians have substantially lower rates of obesity and cancer (4.4% and 0.3% in India compared with 27.9% and 2.8% in England), but higher rates of uncontrolled diabetes, kidney disease, and chronic liver disease (8.9%, 9.7%, and 5.3% in India and 2.1%, 5.6%, and 2.6% in England).

**Table 1.** Prevalence of COVID-19 risk factors in India and England.

|  | Prevalence (%) |  |
| --- | --- | --- |
|  | <u>India</u> | <u>England</u> |
| Age 18-39 | 50.2 | 36.6 |
| Age 40-49 | 19.2 | 16.3 |
| Age 50-59 | 14.3 | 17.0 |
| Age 60-69 | 10.3 | 13.3 |
| Age 70-79 | 4.6 | 10.4 |
| Age 80-99 | 1.5 | 6.3 |
| Male | 47.1 | 48.9 |
| Diabetes (Controlled) | 1.7 | 6.4 |
| Diabetes (Uncontrolled) | 8.9 | 2.1 |
| Hypertension | 28.2 | 28.0 |
| Obese (class I & II) | 4.0 | 24.8 |
| Obese (class III) | 0.4 | 3.1 |
| Chronic Heart Disease | 4.4 | 5.9 |
| Chronic Respiratory Disease | 4.8 | 2.5 |
| Asthma | 2.5 | 9.2 |
| Kidney Disease | 9.7 | 5.6 |
| Chronic Liver Disease | 5.3 | 2.6 |
| Haematological Cancer | 0.0 | 0.2 |
| Non-haematological Cancer | 0.3 | 2.6 |
| Stroke, Dementia | 1.3 | 1.5 |
| Other Neurological Condition | 0.0 | 0.1 |
| Psoriasis, Rheumatoid | 1.0 | 2.4 |
| Other Immunosuppressive Conditions | 0.1 | 0.1 |

We show age-specific prevalence differences between India and England for the conditions for which we have biomarkers in India and are more precisely estimated (figure 1), as well as age-specific prevalence of all conditions for both countries (appendix p 3). Overall rates of diabetes are higher in India at all ages, but diabetes in India is overwhelmingly uncontrolled, while three quarters of diabetes is controlled in England. Hypertension (the sum of controlled and uncontrolled) is higher in India at young ages (31.3% for ages 40–49 in India and 18.3% in England) but lower at higher ages (52.3% at ages 70–79 in India and 61.3% in England) and is overwhelmingly uncontrolled. Conversely, obesity rates are higher at all ages in England.

**Figure 1.**
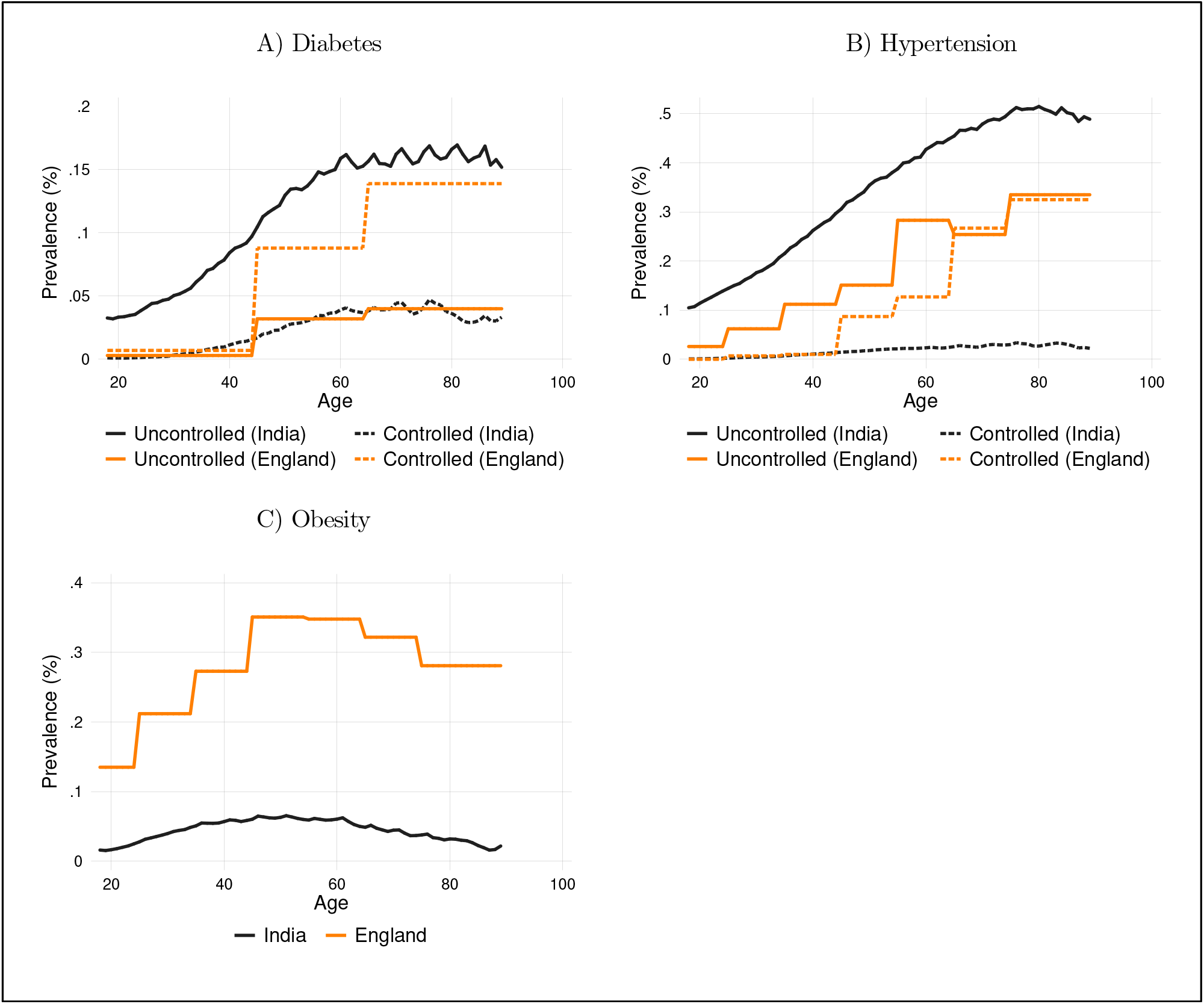
Prevalence of diabetes, hypertension, and obesity in India and England.

### Relative risk of COVID-19 mortality from combined risk factors in India and England

The age-specific population relative risk across all health conditions combined (*PRR*_*a*_) is higher in India than in England at nearly all ages, but the difference in *PRR*_*a*_ between the two countries is below 15% at every age (figure 2). Modelled age-specific mortality rates in India are highest relative to England between ages 40 and 80.

**Figure 2.**
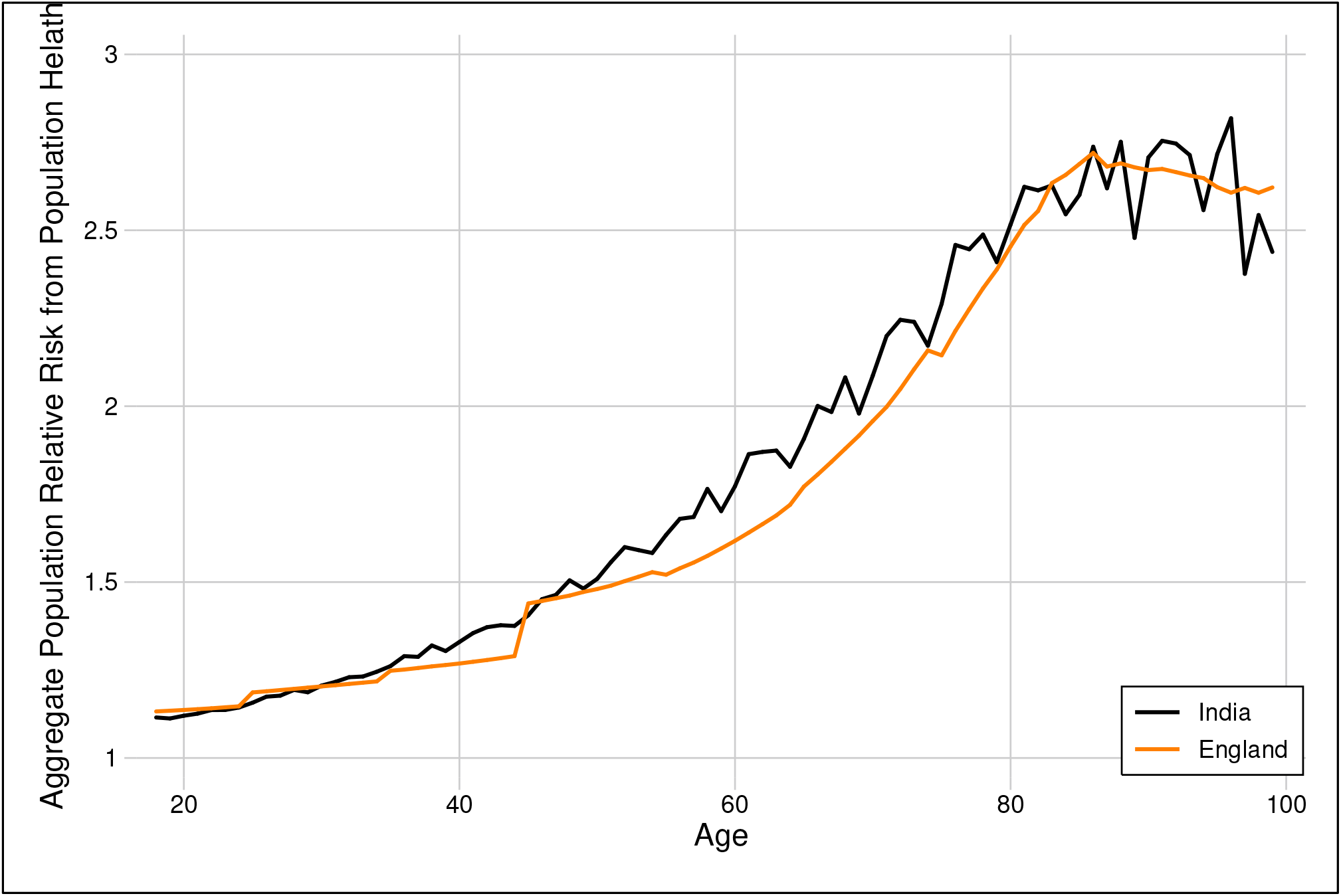
Age-specific population relative risk from combined health conditions (*PRR*_*a*_)

The *PRR*_*a*_ reflects the age-specific prevalence and associated risk of each health condition. Taking risk, prevalence, and population at every age into account provides the full population relative risk of each health condition(*PRR*_*c*_) — the proportional increase in population mortality across all ages driven by each health condition (table 2).

**Table 2.** Population relative risk of COVID-19 mortality from each health condition.

|  | Individual<br>Relative Risk | Population<br>Relative Risk |  |
| --- | --- | --- | --- |
|  |  | <u>India</u> | <u>England</u> |
| Diabetes (Controlled) | 1.50 | 1.007 | 1.032 |
| Diabetes (Uncontrolled) | 2.34 | 1.111 | 1.028 |
| Hypertension | 0.95 | 0.987 | 0.986 |
| Obese (class I & II) | 1.35 | 1.014 | 1.086 |
| Obese (class III) | 2.26 | 1.005 | 1.039 |
| Chronic Heart Disease | 1.27 | 1.013 | 1.033 |
| Chronic Respiratory Disease | 1.77 | 1.043 | 1.019 |
| Asthma | 1.25 | 1.006 | 1.022 |
| Kidney Disease | 1.71 | 1.086 | 1.078 |
| Chronic Liver Disease | 1.61 | 1.034 | 1.020 |
| Haematological Cancer | 3.48 | 1.001 | 1.010 |
| Non-haematological Cancer | 1.56 | 1.002 | 1.026 |
| Stroke, Dementia | 1.78 | 1.011 | 1.028 |
| Other Neurological Conditions | 2.44 | 1.002 | 1.002 |
| Psoriasis, Rheumatoid | 1.23 | 1.003 | 1.008 |
| Other Immunosuppressive Conditions | 1.68 | 1.001 | 1.002 |

Uncontrolled diabetes, which is associated with substantial mortality risk (*RR*_*c*_ 2.34), increases total mortality by 11% in India (*PRR*_*c*_ 1.11), but only 3% in England (*PRR*_*c*_ 1.03), reflecting its significantly higher prevalence in India at all ages. In contrast, controlled diabetes, more common in England than India, raises mortality by 3% in England and only 1% in India (*PRR*_c_ 1.03 vs 1.01). In addition to uncontrolled diabetes, the health conditions causing the largest increases in mortality in India are kidney disease (*PRR*_*c*_ = 1.09), and chronic respiratory disease (*PRR*_*c*_ = 1.04). In England, the most consequential health conditions are obesity (combined *PRR*_*c*_ across all obesity classes = 1.13), and kidney disease (*PRR*_*c*_ = 1.08).

Comparing the percentage difference between the *PRR*_*c*_ of each health condition between India and England (figure 3), the condition with the largest differential impact on mortality between the two countries is uncontrolled diabetes, which increases population mortality by 8.03% in India relative to England. Mortality in India relative to England is also increased by chronic respiratory disease (+2.3%) and chronic liver disease (+1.4%), but decreased by the differential prevalence of obesity (combined - 9.7%), cancer (−3.2%), and controlled diabetes (−2.4%). No other risk factor has an effect of greater than ±2.5% on India’s relative mortality. The combined effect of health conditions leads to 9.7% *higher* mortality in England than in India, reflecting England’s higher age-specific prevalence of certain conditions like obesity and cancer, as well as its older age structure that increases population share at ages with worse health. This differential mortality risk does *not* include the direct effect of older age, which is associated with substantial risk (*RR*_*c*_ 4.68 for age 70-80, 11.92 above 80) and magnifies England’s mortality disadvantage substantially.

**Figure 3.**
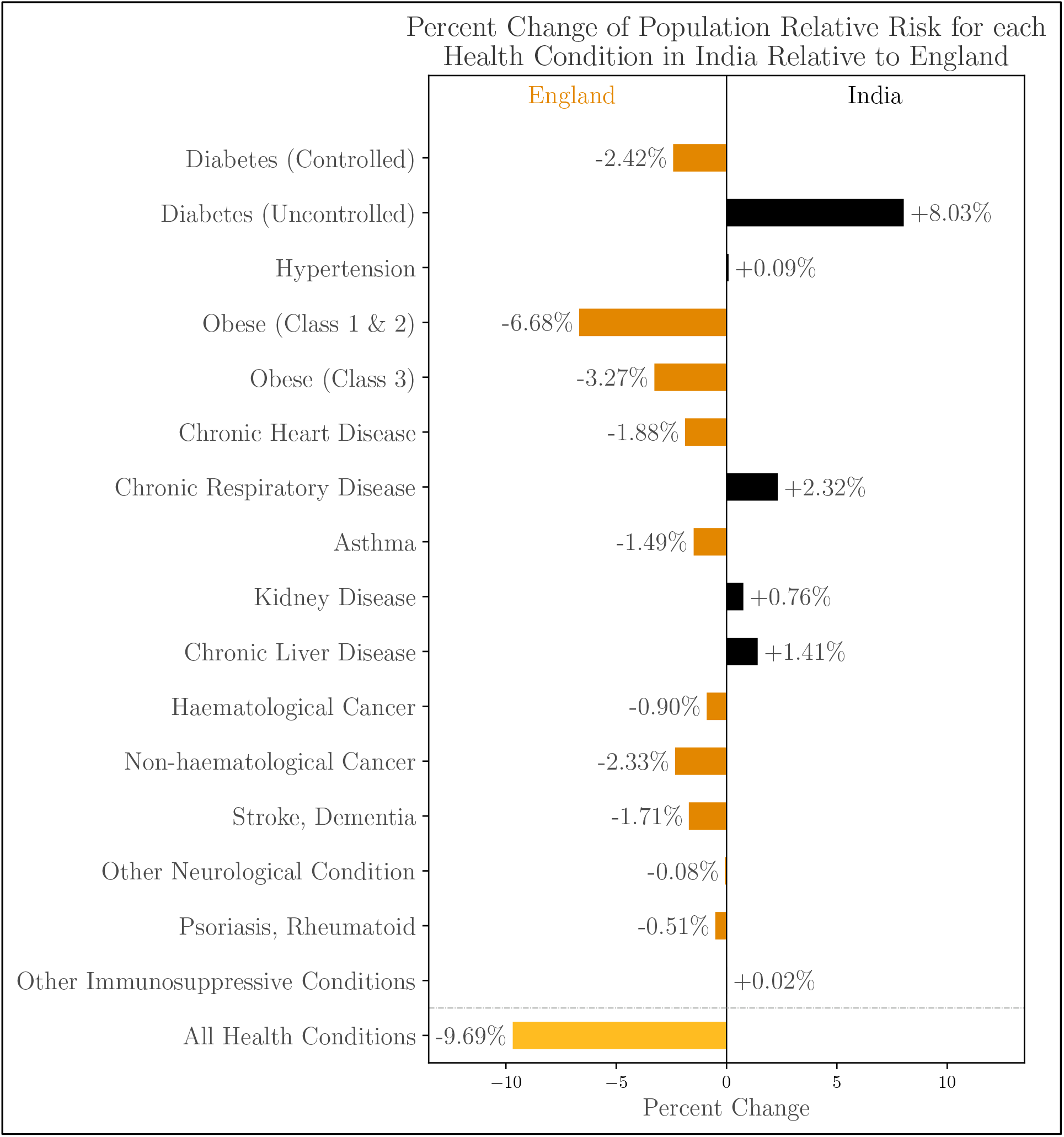
Condition-specific population relative risk in India v. England.

Combining the population relative risk from health conditions with the direct effect of demographics on mortality, we modelled the density of deaths across the age distribution (figure 4). In England, 8.8% of expected deaths are below age 60, closely matching the 6.5% observed in England through May 2020 and the 8.6% reported in the OpenSAFELY dataset. In India, 22.0% of modelled deaths are below age 60, which is substantially lower than the 50% observed in case reports. Applying England’s age-specific prevalence of health conditions to India’s demographic distribution, in order to isolate the effect of health conditions from demographics, results in a distribution nearly identical to the India model. In other words, differences in health conditions between India and England have almost no effect on mortality, indicating that the modelled shift toward younger populations comes from the demographic distribution alone.

**Figure 4.**
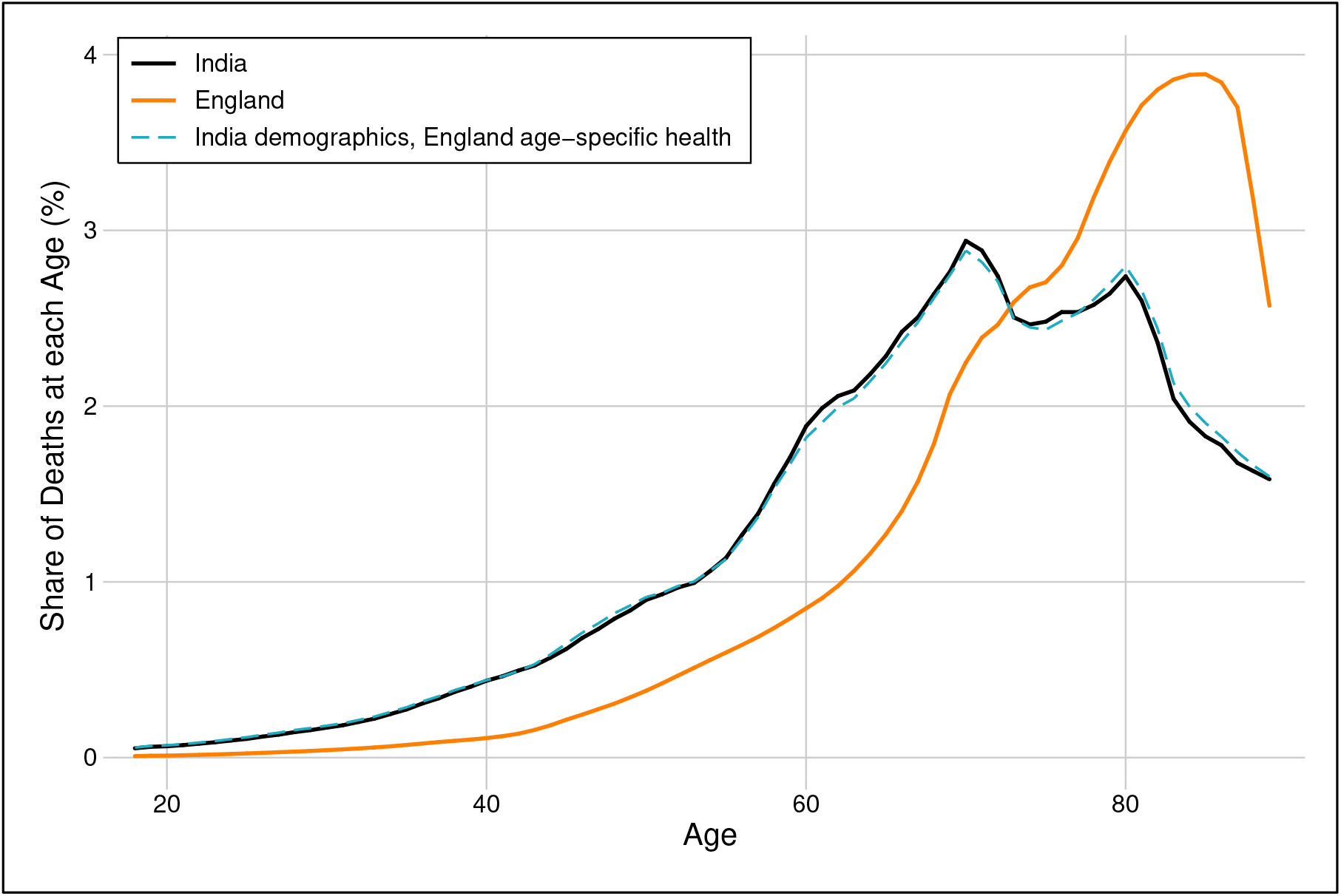
Modelled age distribution of COVID-19 mortality.

In the appendix, we test sensitivity to uncertainty in prevalences and HRs (appendix pp 6–8). The latter estimates cover alternate hazard ratios estimated from other studies.^18,26^ We also test sensitivity to alternate assumptions about covariance of health conditions (appendix p 9). In all cases, we find that the population relative risk from health conditions in England is greater than in India, and that accounting for health conditions cannot explain any of the higher incidence of mortality among the young in India relative to England.

## DISCUSSION

We used the best publicly available data on population health to examine the extent to which demographics and pre-existing health conditions known to increase COVID-19 mortality can account for the disproportionately high share of COVID-19 deaths in younger populations observed in India relative to England. We show that differences in population health do not significantly shift the relative age distribution of disease severity and slightly *lower* aggregate mortality in India relative to England. Higher prevalence of diabetes and respiratory illness raise mortality risk relative to England, but these effects are offset by lower rates of obesity, heart disease, and cancer. While the Indian age distribution substantially shifts expected mortality toward the young, it explains less than a third of the difference in the share of deaths under 60 compared with England.

Epidemiologic models have typically assumed that comorbidities will exacerbate the mortality of COVID-19 in India and other poorer countries relative to rich countries. We found that comorbidities identified as key risk factors in rich countries do not increase expected mortality in India relative to England, in aggregate or among the young. This suggests that understanding the other factors that may explain the differential mortality among the young observed in lower income contexts, such as different patterns of infection, under-resourced health systems, or comorbidities unique to LMICs, should be a priority for further research.

This study improves upon prior work by examining the extent to which comorbidities can explain the younger incidence of COVID-19 mortality in LMICs, by estimating mortality effects of specific comorbidities, and by calibrating a model with a comprehensive set of comorbidity hazard ratios drawn from a large-sample multivariate analysis of COVID-19 mortality. Models calibrated with bivariate hazard ratios or raw prevalences of comorbidities among severe cases are likely to overstate the effect of pre-existing health conditions because of the significant increase in all comorbidities with age alongside the direct effect of age on COVID-19 mortality.

The key limitation of this study is that there are virtually no data on the COVID-19 mortality risks associated with health conditions that are more common in LMICs than in high income countries, such as protein calorie malnutrition, micronutrient deficiency, and HIV/AIDS.^4^ If these conditions make individuals more susceptible to severe infections, then population health may indeed exacerbate the severity of COVID-19 in LMICs. Understanding the extent to which health conditions endemic to poor countries affect COVID-19 severity is an urgent priority, particularly as policy responses increasingly focus on identifying and isolating high risk individuals.^27^

Our analysis is also constrained by the limited and changing evidence on risk factors for COVID-19 severity. Based on the availability of existing measures, our model assumed that health condition relative risks are age-invariant. However, data from New York’s epidemiological surveillance system suggest that hypertension and diabetes may contribute more to mortality at younger ages,^28^ which would exacerbate the burden of illness among the young in LMICs. Further, if illness severity and the quality of prior medical management of pre-existing health conditions change mortality risk for the same diagnosis across contexts, applying HRs from England may understate mortality risk in India. Finally, hazard ratios which are not conditioned on infection may reflect infection risk in addition to disease severity risk and thus may not translate directly to the Indian context.

Recognizing these limitations, we have posted our analysis on an open web platform, allowing estimates to be calibrated with different risk factors, hazard ratios, and data from other countries, as more research on the virus emerges.

## Data Availability

All modelling data and code are available on the public github site https://github.com/devdatalab/covid/. Microdata from India can be obtained from the International Institute for Population Sciences in Mumbai.

## Data Sharing Statement

All aggregate data and code will be made available in a public repository hosted at github.com and a web platform will replicate the analysis and allow use of alternate assumptions or data. Deidentified Indian health microdata is available from the International Institute for Population Sciences in Mumbai.

## Ethics approval

This study was ruled exempt from human subjects review by the Institutional Review Board at Dartmouth College.

## Transparency statement

The lead author affirms that the manuscript is an honest, accurate, and transparent account of the study being reported. No important aspects of the study have been omitted and there are no substantial discrepancies from the study as originally planned.

## Role of the funding source

Asher and Novosad received funding from Emergent Ventures.

## Author Statements

All authors had full access to all the data in the study and shared the final responsibility for the decision to submit for publication. All authors saw and approved the final version of the manuscript.

## Author contributions

The corresponding author attests that all listed authors meet authorship criteria and that no others meeting the criteria have been omitted.

## Copyright/license for publication

*The Corresponding Author has the right to grant on behalf of all authors and does grant on behalf of all authors, a worldwide licence to the Publishers and its licensees in perpetuity, in all forms, formats and media (whether known now or created in the future), to i) publish, reproduce, distribute, display and store the Contribution, ii) translate the Contribution into other languages, create adaptations, reprints, include within collections and create summaries, extracts and/or, abstracts of the Contribution, iii) create any other derivative work(s) based on the Contribution, iv) to exploit all subsidiary rights in the Contribution, v) the inclusion of electronic links from the Contribution to third party material where-ever it may be located; and, vi) licence any third party to do any or all of the above*.

## Competing interests

*All authors have completed the ICMJE uniform disclosure form at www.icmje.org/coi_disclosure.pdf and declare: Asher and Novosad had financial support from Emerging Ventures for the submitted work; no financial relationships with any organisations that might have an interest in the submitted work in the previous three years; no other relationships or activities that could appear to have influenced the submitted work*.

## Notes

### Competing Interest Statement

The authors have declared no competing interest.

### Funding Statement

Asher and Novosad are funded by Emergent Ventures through the Development Data Lab. The funder had no role in study design, data collection, data analysis, data interpretation, or writing.

### Author Declarations

The IRB at Dartmouth College determined that this study does not meet the definition of human subjects research and that IRB review is therefore not required.

